# Clinical Outcome of Asymptomatic COVID-19 Infection Among a Large Nationwide Cohort of 5,621 Hospitalized Patients in Korea

**DOI:** 10.1101/2020.10.25.20218982

**Authors:** Hayne Cho Park, Do Hyoung Kim, Ajin Cho, Juhee Kim, Kyu-sang Yun, Jinseog Kim, Young-Ki Lee

## Abstract

We investigated clinical outcome of asymptomatic coronavirus disease 2019 (COVID-19) and identified risk factors associated with high patient mortality using Korean nationwide public database of 5,621 hospitalized patients. The mortality rate and admission rate to intensive care unit were compared between asymptomatic and symptomatic patients. The prediction model for patient mortality was developed through risk factor analysis among asymptomatic patients. The prevalence of asymptomatic COVID-19 infection was 25.8%. The mortality rates were not different between groups (3.3% vs. 4.5%, p=0.17). However, symptomatic patients were more likely to receive ICU care compared to asymptomatic patients (4.1% vs. 1.0%, p<0.0001). The age-adjusted Charlson comorbidity index score (CCIS) was the most potent predictor for patient mortality in asymptomatic patients. The clinicians should predict the risk of death by evaluating age and comorbidities but not the presence of symptoms.

**Article Summary Line:** Since asymptomatic patients have similar mortality rate with symptomatic patients, the clinicians should not classify clinical severity according to initial presence of symptom.

## Text

Since coronavirus disease 2019 (COVID-19) has been first reported in Wuhan, China in December 2019, it has been spread throughout the world with high infectivity. By August 16, 2020, over 1.8 million new COVID-19 cases and 39,000 new deaths were reported, and the observed case-fatality ratio has been reported up to 13.9 (*1,2*).The diagnosis of COVID-19 is based upon detection of nucleic acid of the severe acute respiratory syndrome coronavirus 2 (SARS-CoV-2) in patient samples by reverse transcriptase-polymerase chain reaction (RT-PCR) whether or not patients have clinical symptoms (*3*). Therefore, COVID-19 infection includes both symptomatic and asymptomatic infections.

The incidence of asymptomatic infections has been varied because screening policies are different across countries (*4*). The difference in the incidence can be due to low awareness of asymptomatic infections in the early outbreak and limited detection capacity in some countries. Recently, the Centers for Disease Control and Prevention announced the revised guideline of testing for COVID-19. The guideline recommended COVID-19 testing for all symptomatic patients while limited testing for asymptomatic patients (*5*). However, asymptomatic infections are known to have the same infectivity as symptomatic infections (*6*). A recent paper demonstrated that viral load in asymptomatic patients were similar to that in symptomatic patients (*7*). Therefore, early diagnosis of asymptomatic infections can be important to prevent further transmission of the viral disease.

Recent papers about asymptomatic patients with COVID-19 demonstrated that their clinical course is mild and most of them did not develop symptoms throughout isolation period (*8,9*). However, there is no study about prognosis of initially asymptomatic carriers upon severe clinical outcome and mortality compared to symptomatic patients. Since Korean government not only performed confirmatory tests for those who are suspected to have the disease but also implemented screening tests for those without symptoms, asymptomatic and mild symptomatic cases were diagnosed in a large proportion. This study was performed among hospitalized patients during COVID-19 outbreak in Korea in order to demonstrate 1) the difference in incidence of severe clinical outcome (mortality and admission to intensive care unit (ICU)) among asymptomatic patients compared to symptomatic patients and 2) risk factors associated with severe outcome among asymptomatic patients.

## Materials and Methods

### Study Design and Participants

This is a retrospective cohort study using the nationwide COVID-19 database provided by Korean Center for Disease Control (KCDC). The database contains the clinical and epidemiologic data as well as clinical outcome of 5,628 confirmed cases who had received treatment from 100 hospitals until April 30, 2020. Among them, 7 patients who were confirmed for COVID-19 after death were excluded, and a total of 5,621 patients were included in the analysis. According to the definition provided by the KCDC (*10*), the confirmed case was defined as a patient who had been confirmed to be infected with COVID-19 by RT-PCR assay or virus isolation from nasal and/or pharyngeal swab specimens regardless of the clinical symptoms. The present study protocol was reviewed and approved by the Institutional Review Board of the Kangnam Sacred Heart Hospital, Seoul, Korea (HKS 2020-06-025). The informed consent was waived due to retrospective nature of the study.

### Definition

Asymptomatic patients were defined as those without any symptoms at admission no matter what they developed symptoms during clinical courses. Symptomatic patients were defined as those who presented symptoms upon admission. The clinical symptoms included febrile sense, cough, sputum, sore throat, rhinorrhea, myalgia, shortness of breath, headache, altered consciousness/confusion, nausea/vomiting, and diarrhea. Anemia was defined as a plasma hemoglobin (Hb) level of less than 12.0 g/dl. Lymphocytopenia was defined as a lymphocyte count of less than 800/μl cells, and thrombocytopenia was defined as a platelet count of less than 150,000/μl.

### Study outcomes

The primary outcome was defined as the patient mortality. The secondary outcome was the rate of ICU admission. Risk factors associated with outcomes were analyzed and compared between asymptomatic and symptomatic patients. We established a prediction model for patient mortality through risk factor analysis among asymptomatic patients.

### Data collection

Demographic data including age by decade, sex, survival status, ICU admission status, and duration of isolation were collected. The body mass index (BMI), systemic and diastolic blood pressure (BP), heart rate, and body temperature at initial visit were also measured and reported. Laboratory assessments consisted of Hb, white blood cell, lymphocyte, and platelet count. The categories of comorbidities were assessed including diabetes mellitus, hypertension, heart failure, chronic heart disease, asthma, chronic obstructive pulmonary disease, chronic kidney disease, malignancy, chronic liver disease, connective tissue disease, and dementia. Age-adjusted Charlson comorbidity index score (CCIS) has been calculated from a weighted index consisted of age and the number and seriousness of comorbid diseases to predict the risk of mortality among COVID-19 patients (*11,12*). Due to lack of available data, all kinds of chronic heart disease were considered to have congestive heart failure and all forms of chronic liver disease were considered to have mild liver disease. The presence of peripheral vascular disease, cerebrovascular disease, peptic ulcer disease, hemiplegia, end-stage renal disease, or AIDS could not be known from the current database. All kinds of malignancy were considered non-metastatic.

### Statistical Analyses

The normally distributed numerical variables were expressed as the mean ± standard deviation, whereas variables with skewed distributions were expressed as the median and interquartile range. Statistical comparisons between continuous variables were performed with an independent Student t-test or one-way analysis of variance (ANOVA) for more than two group. For the data without normal distribution, the Wilcoxon Signed Rank Test for two groups or Kruskal Wallis Test for more than two groups were performed. Categorical measures are presented as percentages. The Chi square test and Fisher’s exact test were applied to categorical variables as appropriate.

The Kaplan-Meier method was used to compare death-free survival curves, and differences were assessed utilizing the log-rank test. We used univariate and multivariate Cox proportional hazard model to estimate risk factors associated with patient mortality. Age was excluded from the multivariate analysis because of its potential interaction with CCIS. We used univariate and multivariate logistic regression models to evaluate the risk factors for ICU admission. A nomogram to predict 14-day and 28-day mortality risk of the patient was built based on the variables found in multivariate Cox proportional hazard model. In the nomogram, CCIS was used as a continuous variable to check the impact per score. The maximum score of each variable was set as 100. The performance of the nomogram was measured based on the Harrell concordance index (C-index). The nomogram was validated in calibration plots with 1,000 bootstrap samples in which the estimated survival probability was compared with the observed value. All statistical analysis was performed by using R version 4.0.2 (R Foundation for Statistical Computing; http://www.r-project.org/). P value <0.05 was considered statistically significant.

## Results

### Comparison of baseline characteristics between asymptomatic and symptomatic patients

In a total of 5,621 patients, asymptomatic patients were 1,449 (25.8%) and symptomatic patients were 4,172 (74.2%). Baseline characteristics were compared between asymptomatic and symptomatic patients (Table 1). In the asymptomatic group, the proportions of patients under the age of 30 (30.0%) and over 70 years of age (17.5%) were greater than those in symptomatic group (22.9% and 14.5%, respectively) (Appendix). A total of 748 patients (51.6%) in asymptomatic group and 2,556 patients (61.3%) in symptomatic group were female. The proportion of patients with low BMI (<18.5 kg/m^2^) was greater in asymptomatic group than that of symptomatic group. Asymptomatic patients had higher prevalence of dementia and lower prevalence of asthma than symptomatic group. In addition, the absolute lymphocyte and platelet counts were significantly lower in symptomatic patients compared to asymptomatic patients. In symptomatic patients, cough (56.1%) was the most common symptom followed by sputum (38.8%), febrile sense (31.2%), headache (23.2%), myalgia (22.2%), sore throat (21.1%), dyspnea (15.9%), rhinorrhea (14.9%), diarrhea (12.4%), nausea or vomiting (5.9%), fatigue (5.6%), and altered mentality (0.8%). The proportions of patients with no comorbidity (CCIS 0, 38.7% vs. 31.2%) and high CCIS (≥5) (15.2% vs. 11.9%) were greater in asymptomatic patients compared to symptomatic patients.

**Table 1.**
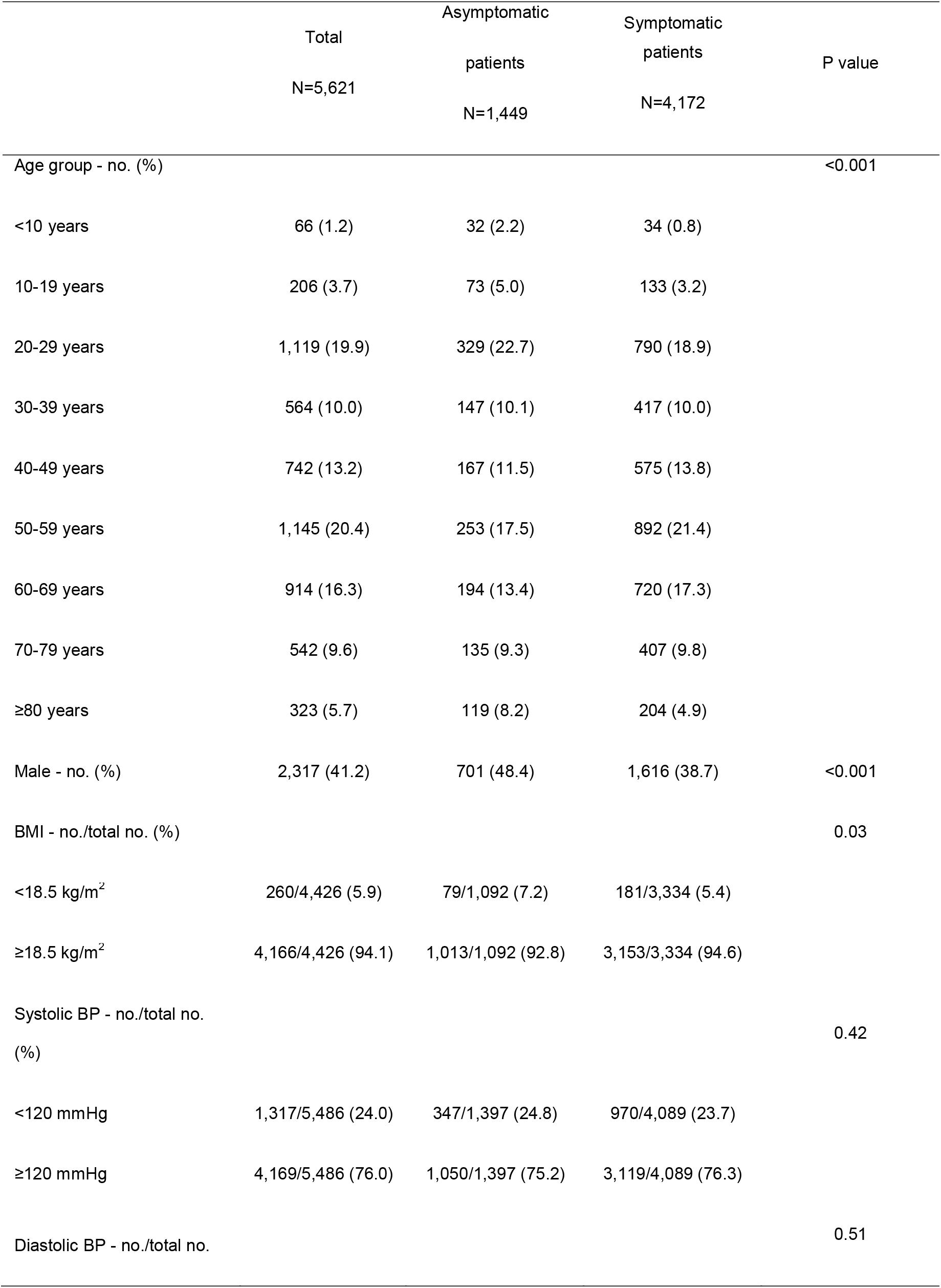
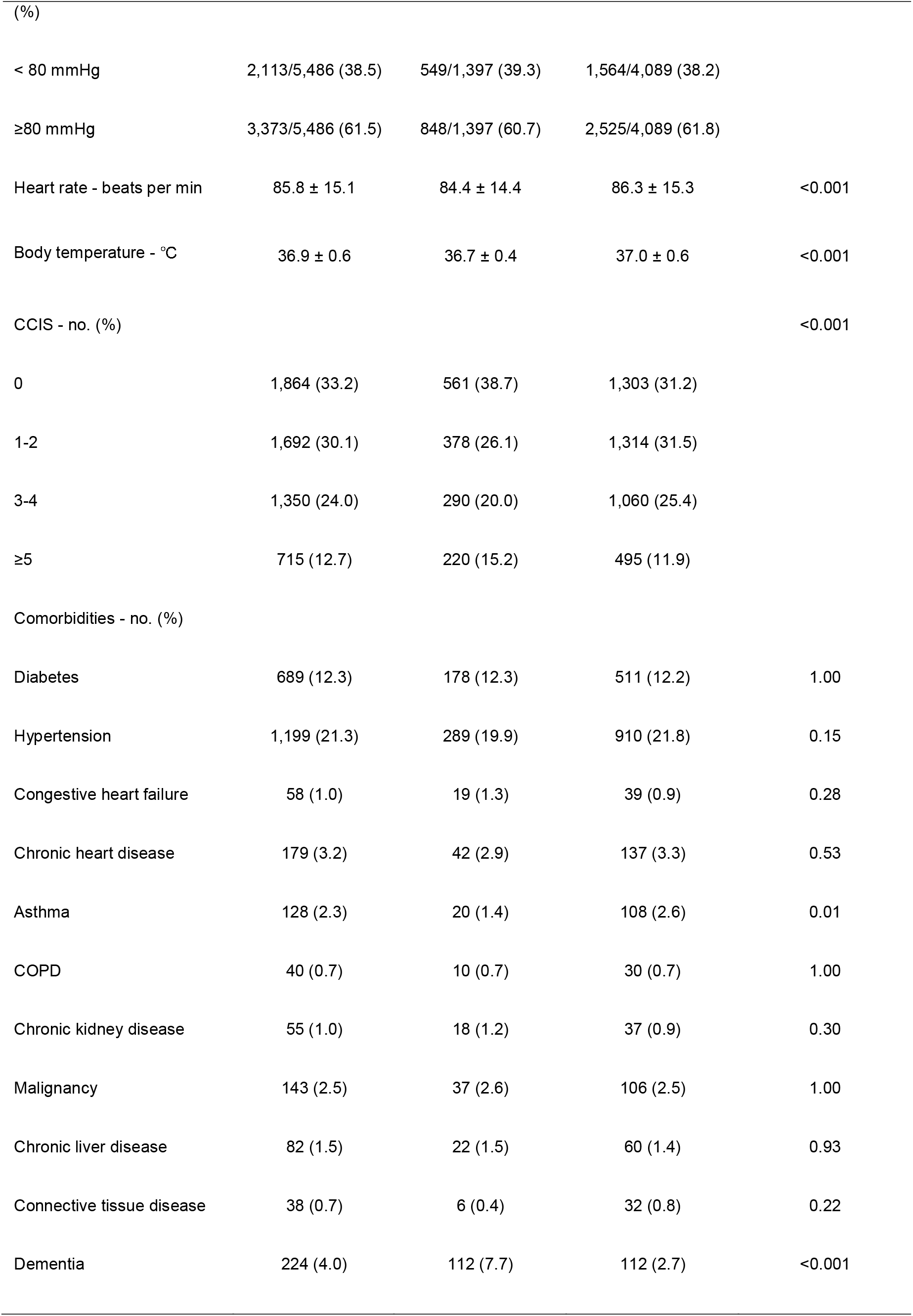
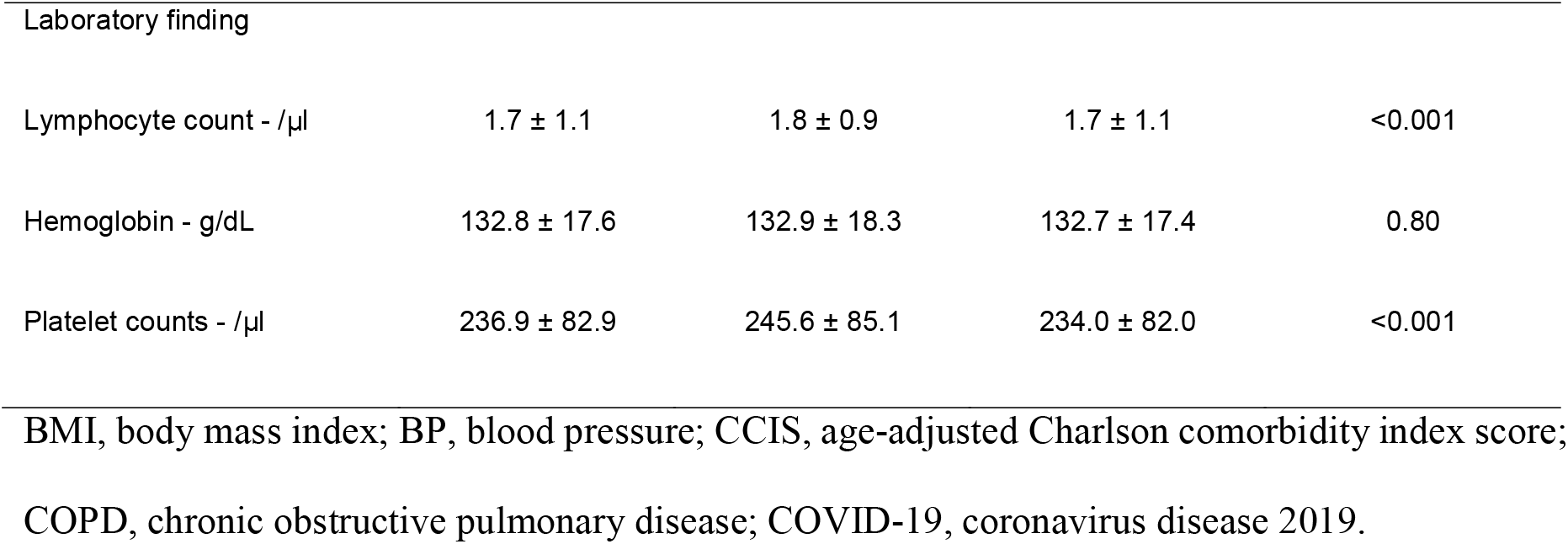
Clinical characteristics between asymptomatic and symptomatic patients with COVID-19

|  | Total<br>N=5,621 | Asymptomatic<br>patients<br>N=1,449 | Symptomatic<br>patients<br>N=4,172 | P value |
| --- | --- | --- | --- | --- |
| Age group - no. (%) |  |  |  | <0.001 |
| <10 years | 66 (1.2) | 32 (2.2) | 34 (0.8) |  |
| 10-19 years | 206 (3.7) | 73 (5.0) | 133 (3.2) |  |
| 20-29 years | 1,119 (19.9) | 329 (22.7) | 790 (18.9) |  |
| 30-39 years | 564 (10.0) | 147 (10.1) | 417 (10.0) |  |
| 40-49 years | 742 (13.2) | 167 (11.5) | 575 (13.8) |  |
| 50-59 years | 1,145 (20.4) | 253 (17.5) | 892 (21.4) |  |
| 60-69 years | 914 (16.3) | 194 (13.4) | 720 (17.3) |  |
| 70-79 years | 542 (9.6) | 135 (9.3) | 407 (9.8) |  |
| ≥80 years | 323 (5.7) | 119 (8.2) | 204 (4.9) |  |
| Male - no. (%) | 2,317 (41.2) | 701 (48.4) | 1,616 (38.7) | <0.001 |
| BMI - no./total no. (%) |  |  |  | 0.03 |
| <18.5 kg/m<sup>2</sup> | 260/4,426 (5.9) | 79/1,092 (7.2) | 181/3,334 (5.4) |  |
| ≥18.5 kg/m<sup>2</sup> | 4,166/4,426 (94.1) | 1,013/1,092 (92.8) | 3,153/3,334 (94.6) |  |
| Systolic BP - no./total no.<br>(%) |  |  |  | 0.42 |
| <120 mmHg | 1,317/5,486 (24.0) | 347/1,397 (24.8) | 970/4,089 (23.7) |  |
| ≥120 mmHg | 4,169/5,486 (76.0) | 1,050/1,397 (75.2) | 3,119/4,089 (76.3) |  |
| Diastolic BP - no./total no. |  |  |  | 0.51 |
| (%) |  |  |  |  |
| < 80 mmHg | 2,113/5,486 (38.5) | 549/1,397 (39.3) | 1,564/4,089 (38.2) |  |
| ≥80 mmHg | 3,373/5,486 (61.5) | 848/1,397 (60.7) | 2,525/4,089 (61.8) |  |
| Heart rate - beats per min | 85.8 ± 15.1 | 84.4 ± 14.4 | 86.3 ± 15.3 | <0.001 |
| Body temperature - °C | 36.9 ± 0.6 | 36.7 ± 0.4 | 37.0 ± 0.6 | <0.001 |
| CCIS - no. (%) |  |  |  | <0.001 |
| 0 | 1,864 (33.2) | 561 (38.7) | 1,303 (31.2) |  |
| 1-2 | 1,692 (30.1) | 378 (26.1) | 1,314 (31.5) |  |
| 3-4 | 1,350 (24.0) | 290 (20.0) | 1,060 (25.4) |  |
| ≥5 | 715 (12.7) | 220 (15.2) | 495 (11.9) |  |
| Comorbidities - no. (%) |  |  |  |  |
| Diabetes | 689 (12.3) | 178 (12.3) | 511 (12.2) | 1.00 |
| Hypertension | 1,199 (21.3) | 289 (19.9) | 910 (21.8) | 0.15 |
| Congestive heart failure | 58 (1.0) | 19 (1.3) | 39 (0.9) | 0.28 |
| Chronic heart disease | 179 (3.2) | 42 (2.9) | 137 (3.3) | 0.53 |
| Asthma | 128 (2.3) | 20 (1.4) | 108 (2.6) | 0.01 |
| COPD | 40 (0.7) | 10 (0.7) | 30 (0.7) | 1.00 |
| Chronic kidney disease | 55 (1.0) | 18 (1.2) | 37 (0.9) | 0.30 |
| Malignancy | 143 (2.5) | 37 (2.6) | 106 (2.5) | 1.00 |
| Chronic liver disease | 82 (1.5) | 22 (1.5) | 60 (1.4) | 0.93 |
| Connective tissue disease | 38 (0.7) | 6 (0.4) | 32 (0.8) | 0.22 |
| Dementia | 224 (4.0) | 112 (7.7) | 112 (2.7) | <0.001 |
| Laboratory finding |  |  |  |  |
| Lymphocyte count - /μl | 1.7 ± 1.1 | 1.8 ± 0.9 | 1.7 ± 1.1 | <0.001 |
| Hemoglobin - g/dL | 132.8 ± 17.6 | 132.9 ± 18.3 | 132.7 ± 17.4 | 0.80 |
| Platelet counts - /μl | 236.9 ± 82.9 | 245.6 ± 85.1 | 234.0 ± 82.0 | <0.001 |

BMI, body mass index; BP, blood pressure; CCIS, age-adjusted Charlson comorbidity index score;
COPD, chronic obstructive pulmonary disease; COVID-19, coronavirus disease 2019.

### Clinical outcomes according to the presence of initial symptoms

Of the 234 patients (4.2 %) who died during hospitalization, 48 (3.3%) were asymptomatic and 186 (4.5%) were symptomatic patients. In the Kaplan-Meier analysis, mortality rate was not statistically different between asymptomatic and symptomatic patients (P=0.17; Figure 1). However, the patients with old age and high CCIS showed higher mortality rate in both symptomatic and asymptomatic groups (Appendix). Symptomatic patients with dementia, malignancy, connective tissue disease and diabetes had higher mortality compared to asymptomatic patients. On the other hands, asymptomatic patients with chronic obstructive pulmonary disease, chronic kidney disease and chronic heart disease showed higher mortality than symptomatic group. Table 2 showed the Cox proportional-hazards models for factors associated with in-hospital death. The male sex, low BMI (<18.5 kg/m^2^), high CCIS (≥3), anemia, lymphocytopenia and thrombocytopenia were associated with high mortality. However, risk of death was not different according to the presence of symptom except dyspnea.

**Table 2.** Predictors associated with mortality in the patients with COVID-19

|  | Univariate |  | Multivariate |  |
| --- | --- | --- | --- | --- |
|  | Hazard ratio (95% CI) | P value | Hazard ratio (95% CI) | P value |
| Age 50-69 years | 14.00 (5.04-38.89) | <0.001 |  |  |
| ≥70 years | 137.37 (51.00-370.03) | <0.001 |  |  |
| Male | 1.63 (1.26-2.11) | <0.001 | 1.55 (1.05-2.29) | 0.03 |
| BMI <18.5 kg/m<sup>2</sup> | 2.50 (1.48-4.22) | <0.001 | 2.88 (1.66-4.99) | <0.001 |
| Systolic BP <120 mmHg | 1.18 (0.87-1.59) | 0.28 |  |  |
| Diastolic BP <80 mmHg | 1.36 (1.04-1.77) | 0.02 | 0.74 (0.50-1.09) | 0.13 |
| Heart rate ≥100 /min | 1.74 (1.29-2.33) | <0.001 | 1.07 (0.68-1.69) | 0.76 |
| Body temperature ≥37.5°C | 2.13 (1.60-2.83) | <0.001 | 1.18 (0.58-2.39) | 0.65 |
| CCIS ≥3 | 61.41 (27.29-138.15) | <0.001 | 22.96 (7.20-73.24) | <0.001 |
| Any symptom | 1.25 (0.91-1.72) | 0.17 |  |  |
| Febrile sense | 2.02 (1.55-2.63) | <0.0001 | 1.31 (0.66-2.62) | 0.44 |
| Fatigue | 1.74 (1.06-2.85) | 0.028 | 1.15 (0.60-2.20) | 0.67 |
| Dyspnea | 6.39 (4.94-8.26) | <0.0001 | 2.90 (1.96-4.29) | <0.0001 |
| Altered mentality | 19.77 (12.46-31.35) | <0.0001 | 2.03 (0.92-4.48) | 0.081 |
| Hemoglobin <12.0 g/dl | 3.99 (3.07-5.19) | <0.001 | 2.08 (1.40-3.11) | <0.001 |
| Lymphocyte counts <800 /μl | 8.36 (6.41-10.89) | <0.001 | 2.61 (1.75-3.89) | <0.001 |
| Platelet counts <150,000 /μl | 3.95 (3.01-5.19) | <0.001 | 2.03 (1.35-3.04) | <0.001 |

BMI, body mass index; BP, blood pressure; CCIS, age-adjusted Charlson comorbidity index score; CI, confidence interval; COVID-19, coronavirus disease 2019; Hb, hemoglobin; HR, hazard ratio.

**Figure 1.**
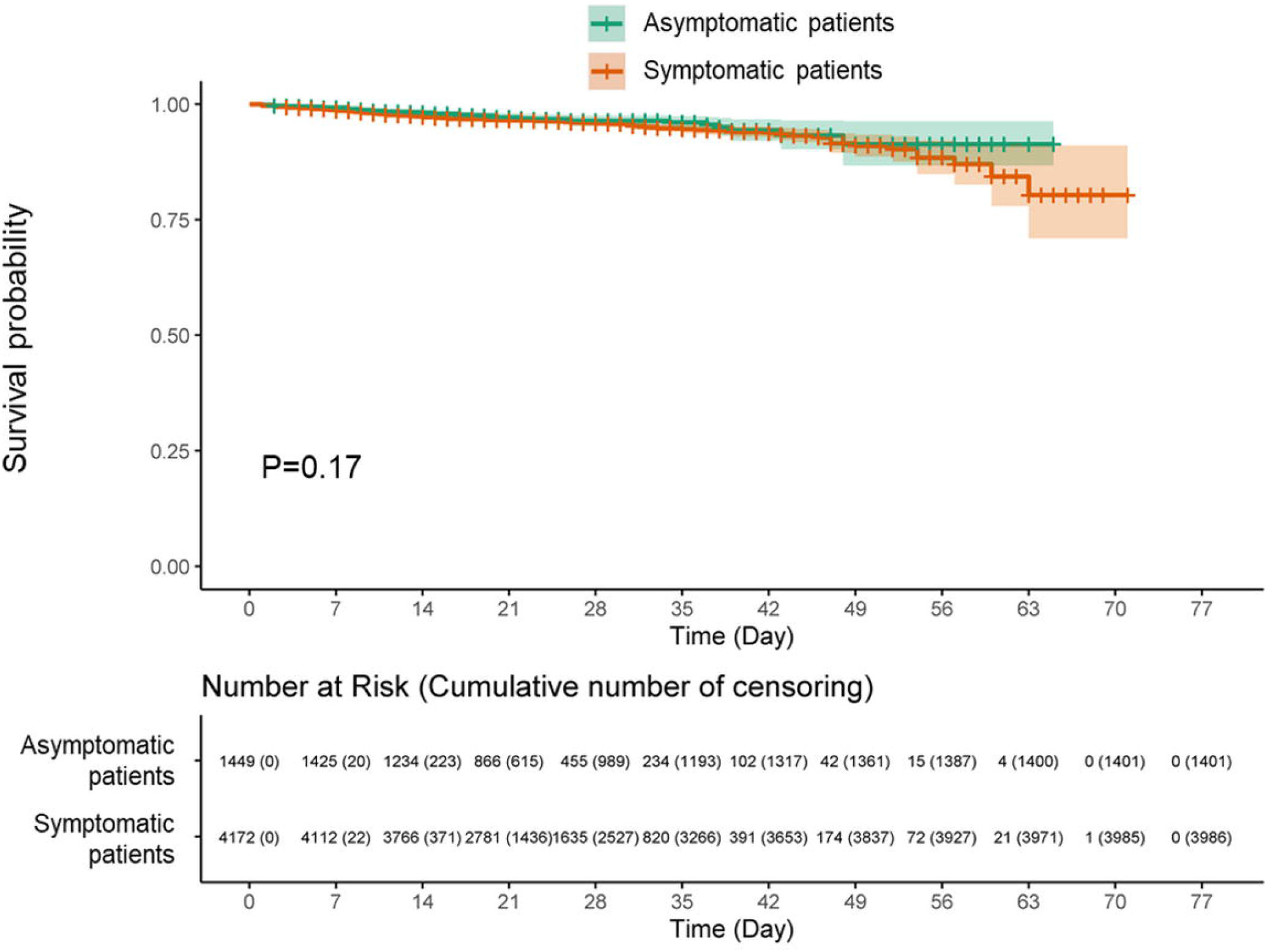
Kaplan-Meier survival plots for mortality according to the presence of symptoms. The figure displays the Kaplan-Meier survival plots of mortality between symptomatic and asymptomatic COVID-19 patients. There was no statistically significant difference between groups (P=0.17). COVID-19, coronavirus disease 2019.

While there was no difference in mortality rate between asymptomatic and symptomatic patients, symptomatic patients were more likely to be admitted to ICU compared to asymptomatic patients. A total of 172 (4.1%) symptomatic patients were admitted to ICU while only 15 (1.0%) asymptomatic patients were during hospitalization. In multivariate analysis, male sex, high CCIS (≥3), anemia, and lymphocytopenia were related to ICU admission (Appendix). In addition, dyspnea (hazard ratio (HR) 4.65, 95% confidence interval (CI) 3.13-6.90) and altered mentality (HR 6.06, 95% CI 1.56-23.50) were risk factors of ICU admission.

### Risk factors for severe clinical outcomes in asymptomatic patients

We assessed the risk factors for mortality in asymptomatic patients. In multivariate analysis, high CCIS (≥5) was an independent risk factor for mortality (HR 61.91, 95% CI 8.29-462.62) together with lymphocytopenia (HR 2.46, 95% CI 1.18-5.11) and thrombocytopenia (HR 2.60, 95% CI 1.31-5.19) (Table 3).

**Table 3.** Cox proportional analysis of predictors associated with mortality in asymptomatic patients with COVID-19

|  | Univariate |  | Multivariate |  |
| --- | --- | --- | --- | --- |
|  | Hazard ratio (95% CI) | P value | Hazard ratio (95% CI) | P value |
| Age ≥70years | 30.98 (13.16-72.94) | <0.001 |  |  |
| Male | 0.69 (0.38-1.22) | 0.20 |  |  |
| BMI <18.5 kg/m<sup>2</sup> | 2.56 (0.56-11.61) | 0.22 |  |  |
| Systolic BP <120 mmHg | 1.23 (0.63-2.39) | 0.54 |  |  |
| Diastolic BP <80 mmHg | 1.12 (0.62-2.03) | 0.71 |  |  |
| Heart rate ≥100 beats per min | 1.13 (0.51-2.54) | 0.76 |  |  |
| CCIS 3-4 | 22.20 (2.73-180.44) | 0.004 | 6.58 (0.73-59.35) | 0.09 |
| ≥5 | 158.80 (21.82-1155.85) | <0.001 | 61.91 (8.29-462.62) | <0.001 |
| Diabetes | 4.54 (2.54-8.10) | <0.001 |  |  |
| Hypertension | 4.99 (2.82-8.83) | <0.001 |  |  |
| Congestive heart failure | 8.79 (3.47-22.27) | <0.001 |  |  |
| Chronic heart disease | 6.64 (3.10-14.19) | <0.001 |  |  |
| Asthma | 2.81 (0.68-11.58) | 0.15 |  |  |
| COPD | 19.71 (7.74-50.22) | <0.001 |  |  |
| Chronic kidney disease | 12.83 (5.43-30.30) | <0.001 |  |  |
| Malignancy | 1.82 (0.44-7.51) | 0.41 |  |  |
| Chronic liver disease | 1.64 (0.23-11.91) | 0.63 |  |  |
| Dementia | 11.15 (6.32-19.69) | <0.001 |  |  |
| Hemoglobin <12.0 g/dl | 5.42 (2.95-9.95) | <0.001 | 1.55 (0.81-2.94) | 0.19 |
| Lymphocyte counts <800 /μl | 5.53 (2.85-10.73) | <0.001 | 2.46 (1.18-5.11) | 0.02 |
| Platelet counts <150,000 /μl | 4.70 (2.53-8.76) | <0.001 | 2.60 (1.31-5.19) | 0.007 |

BMI, body mass index; BP, blood pressure; CCIS, age-adjusted Charlson comorbidity index score; CI, confidence interval; COPD, chronic obstructive pulmonary disease; COVID-19, coronavirus disease 2019; Hb, hemoglobin; HR, hazard ratio.

The predictive nomogram was constructed based on the multivariate Cox analysis for mortality. The 14- and 28-day overall survival probability was calculated from the summed points of CCIS, anemia, lymphocytopenia, and thrombocytopenia (Figure 2). The nomogram showed that CCIS was the most important factor contributing to the prognosis followed by the thrombocytopenia, lymphocytopenia, and anemia. The C-index value for prediction of overall survival was 0.8921, and R^2^ value was 0.992 in 14-day and 0.997 in 28-day prediction model (Appendix).

**Figure 2.**
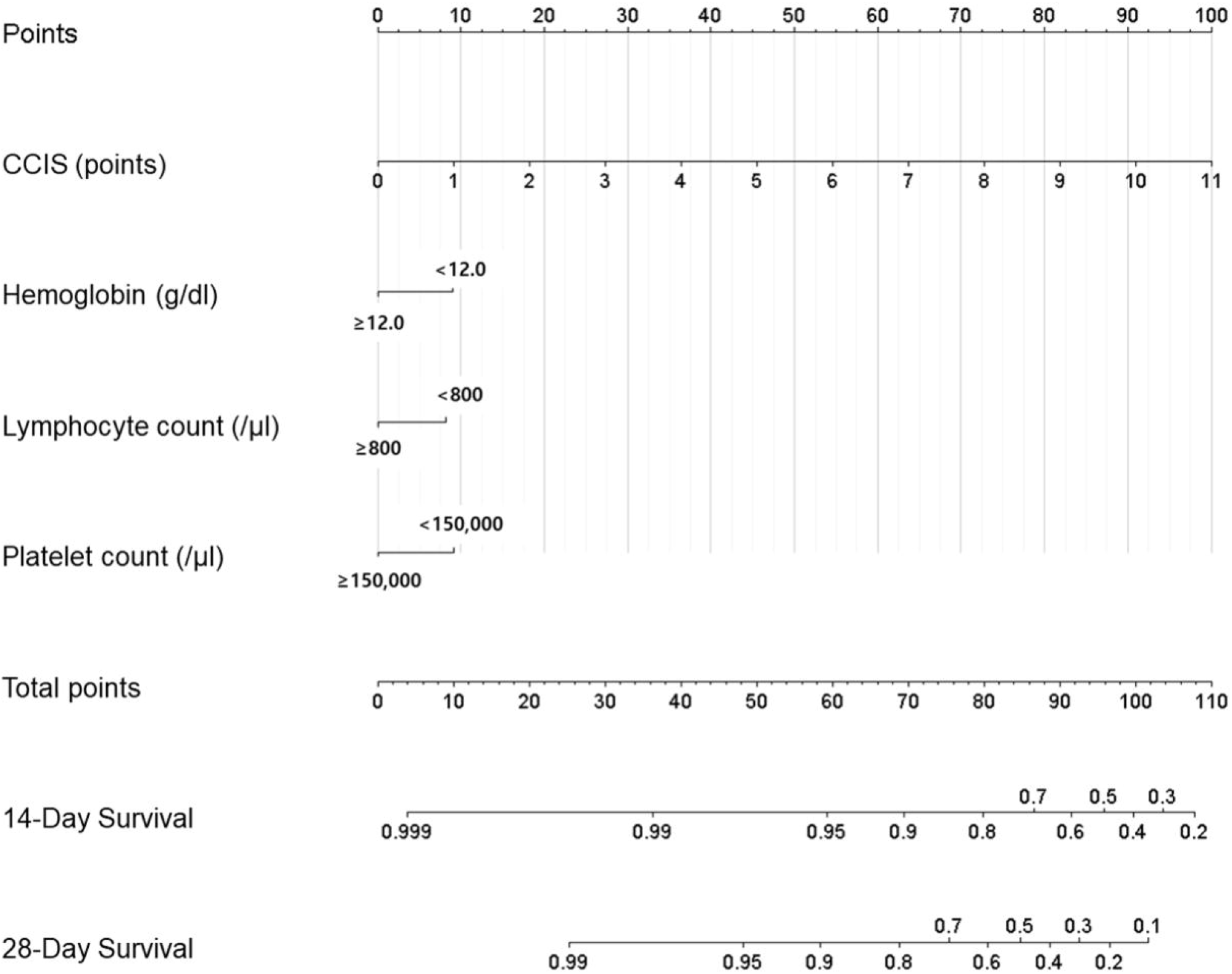
Prognostic nomogram for prediction of the overall survival probability of asymptomatic patients with COVID-19. The nomogram demonstrates CCIS is the potent predictor for 14-day and 28-day survival of the patients. CCIS, age-adjusted Charlson comorbidity index score; COVID-19, coronavirus disease 2019.

## Discussion

This study examined the incidence of mortality and ICU admission among asymptomatic and symptomatic patients during hospitalization due to COVID-19 infection in Korea. The mortality rate in asymptomatic patients was as high as that in symptomatic patients (3.3% vs. 4.5%, P=0.17). However, admission rate to ICU was greater in symptomatic patients compared to that in asymptomatic patients (4.1% vs. 1.0%, P<0.0001). The CCIS was the most potent predictor for mortality in asymptomatic patients.

Previous epidemiologic and observation cohort studies were performed to explain clinical course and outcomes among hospitalized patients (*13-15*). In most of the countries, screening tests for COVID-19 are performed against those who developed symptoms. Recent guideline by the Centers for Disease Control and Prevention also did not recommend the COVID-19 test for asymptomatic patients who are in close contact with the confirmed case. However, there has been no evidence about whether symptomatic and asymptomatic patients should be dealt differently from screening, diagnosis, and treatment. There has been no study comparing clinical outcome between asymptomatic and symptomatic patients.

To our knowledge, this is the first study to evaluate incidence of mortality and its risk factors in asymptomatic patients upon admission. Interestingly, the presence of symptom at diagnosis was not as important as age or comorbidities in prediction of patient mortality. However, symptomatic patients at admission were more likely to receive ICU care. Previous study inferred that patients may spread virus 1 to 3 days before symptom development, and the duration of infectious period may be 6.5 to 9.6 days in asymptomatic patients (*16*). Another recent article demonstrated that the cycle threshold values in asymptomatic patients were similar to those in symptomatic patients, which suggest that asymptomatic patients have similar infectious capacity compared to symptomatic patients (*17*). On the other hands, the clinical severity of COVID-19 may be related to viral load. Recent brief article by Liu et al. demonstrated that the patients with severe COVID-19 tend to have a high viral load and a long viral shedding period compared to the patients with mild COVID-19 (*18*). Another article by German researchers found that the duration of viral shedding was not related to viral replication and isolation from tissue (*19*). Therefore, a complex dynamic between the virus and immune reaction of the host may underlie the severity of the disease and clinical course of COVID-19. In that venue, it is not surprising to find that symptomatic patients were more likely to show severe clinical course and receive ICU care during hospitalization.

However, the mortality rate was not different among asymptomatic and symptomatic patients. Compared to symptomatic patients, asymptomatic patients in our cohort demonstrated greater proportion of elderly population, low BMI, male prevalence, greater proportion of high CCIS (≥5) and higher prevalence of dementia. On the other hands, the degree of lymphocytopenia and thrombocytopenia, which are associated with clinical severity of the disease, were milder in asymptomatic patients. Previous papers well documented the effect of comorbidities or CCIS upon case-fatality rate. In Italy where one of the highest case-fatality rates were reported, the non-survivor demonstrated advanced age and higher CCIS compared to survivors (*13*). Chinese researchers suggested predictive nomogram for fatal outcome including age and preexisting comorbid conditions (*14*). Therefore, the host factors including age and comorbidities are more important in determining the case-fatality rate than the virus factors or the presence and severity of symptoms.

There are some limitations to our study. First, our study excluded those who admitted to the community treatment centers and only included the hospitalized patients. Therefore, there can be a selection bias to generalize our results. Second, we defined asymptomatic patients as those who presented without symptoms at admission. Therefore, some of them would have been developed symptoms at some time after admission. The previous study by Korean researchers demonstrated that about one-third of initially asymptomatic patients developed symptoms during clinical course (*8*). Therefore, we cannot conclude from our study that the prognosis of asymptomatic patients during entire clinical course is similar to that of those who developed symptoms afterwards. Lastly, the patients with dementia and chronic obstructive pulmonary disease or asthma may have under-reported their symptoms, which may result in overestimation of asymptomatic patients.

In conclusion, we found that asymptomatic patients have same mortality risk with symptomatic patients with COVID-19. Our study suggest that asymptomatic patients should not be considered ‘less severe’ than symptomatic patients in treating COVID-19. Regardless of the presence of symptoms, we should predict the clinical risks of the patients based upon age and comorbidities and treat them accordingly.

## Data Availability

Identifiable patient level data from this cohort study are not available to the public.

## Acknowledgments

We acknowledge all the health-care workers involved in the diagnosis and treatment of COVID-19 patients in South Korea. We thank the Central Disease Control Headquarters, National Medical Center and the Health Information Manager in 100 hospitals for their effort in collecting the medical records. There was no specific funding for this work. The authors declare no competing interests. All authors have submitted the ICMJE Form for Disclosure of Potential Conflicts of Interest.

## Author Bio

Dr. Park is an associate professor at the Department of Internal Medicine, Hallym University College of Medicine and Kangnam Sacred Heart Hospital. Her research interests include hereditary kidney disease and critical care medicine.

