## Appendix for "Clinical Outcome of Asymptomatic COVID-19 Infection Among a Large Nationwide Cohort of 5,621 Hospitalized Patients in Korea"

Appendix Figure 1. Demographic distribution of the patients with COVID-19 in Korea. Age distribution according to presence of symptoms. COVID-19, coronavirus disease 2019.

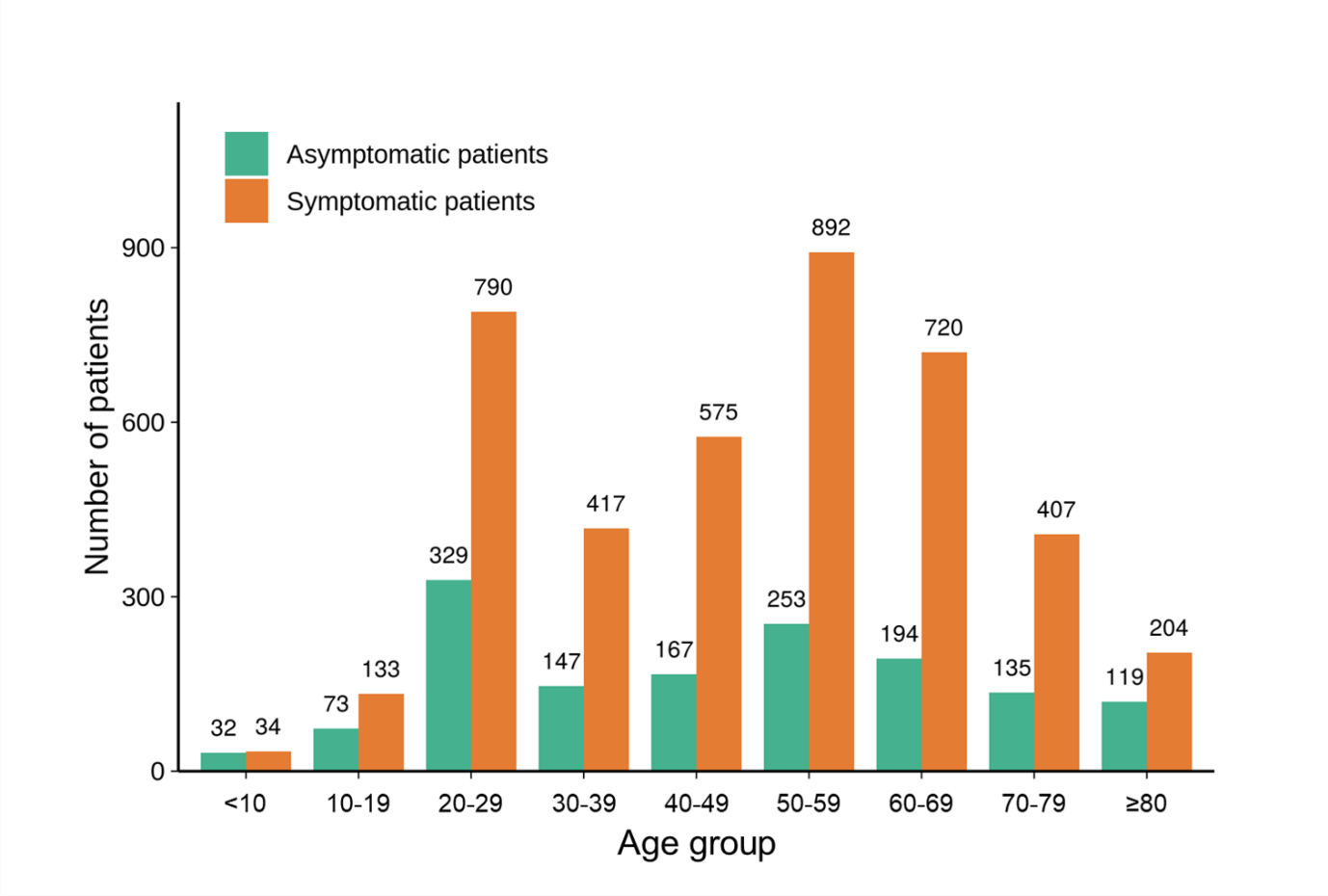

Appendix Figure 2. Percentage of death according to age, CCIS, and comorbidities. The percentage of death in total patients according to (A) age, (B) CCIS, and (C) comorbidities according to presence of symptoms. age-adjusted Charlson comorbidity index score, CHF, Congestive heart failure; CKD, Chronic kidney disease; CLD, Chronic liver disease; COPD, Chronic obstructive pulmonary disease; CTD, Connective tissue disease; HTN, hypertension.

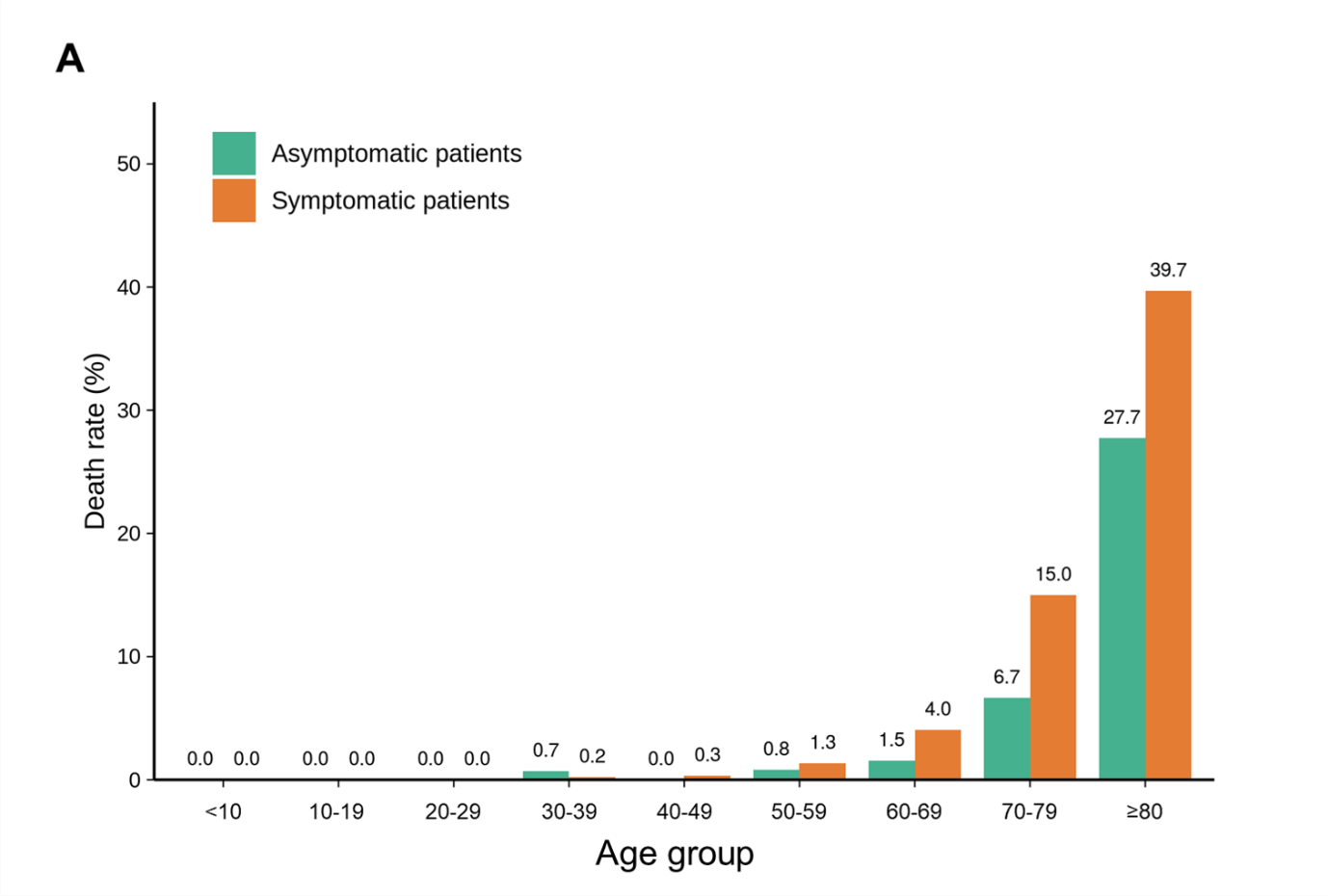

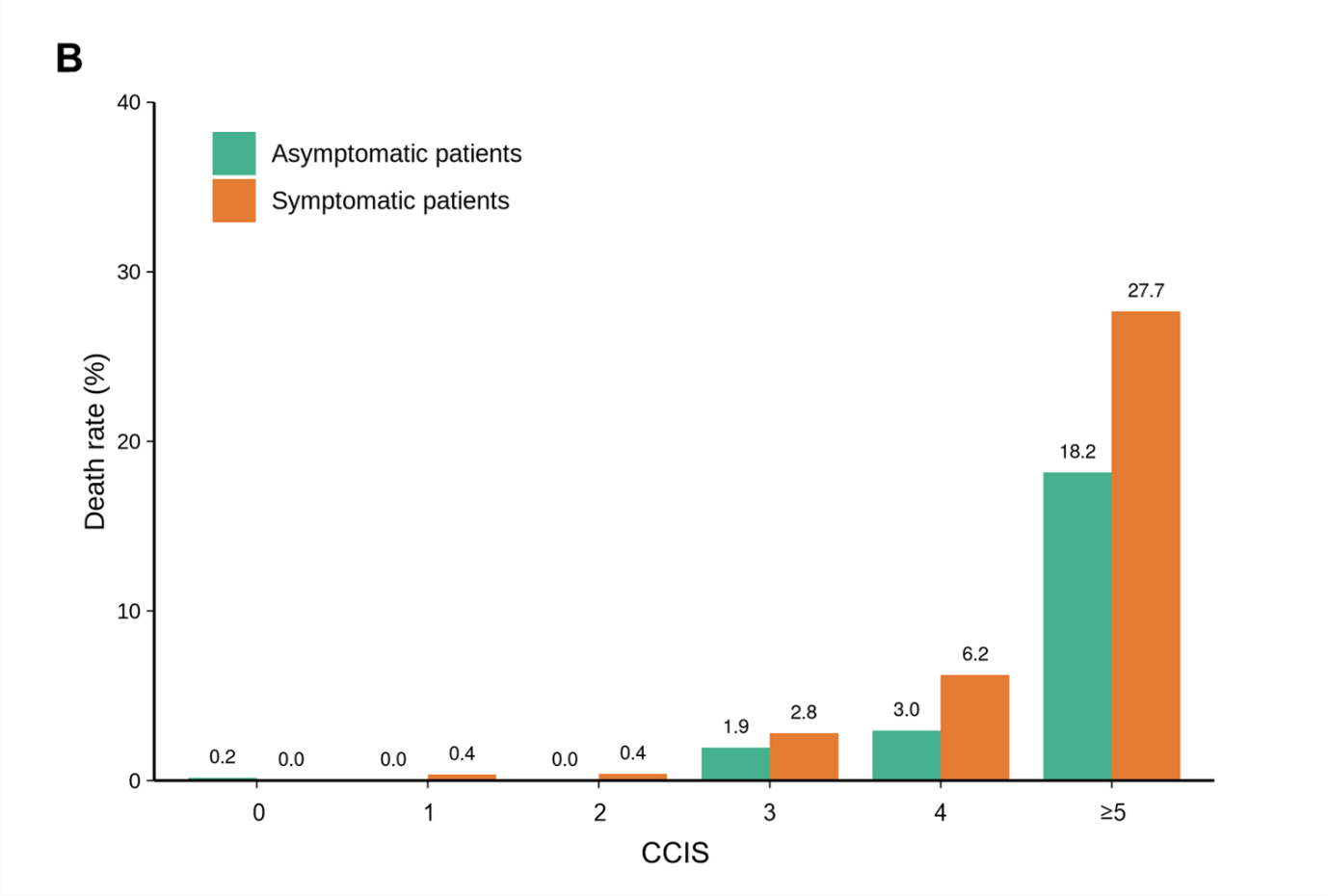

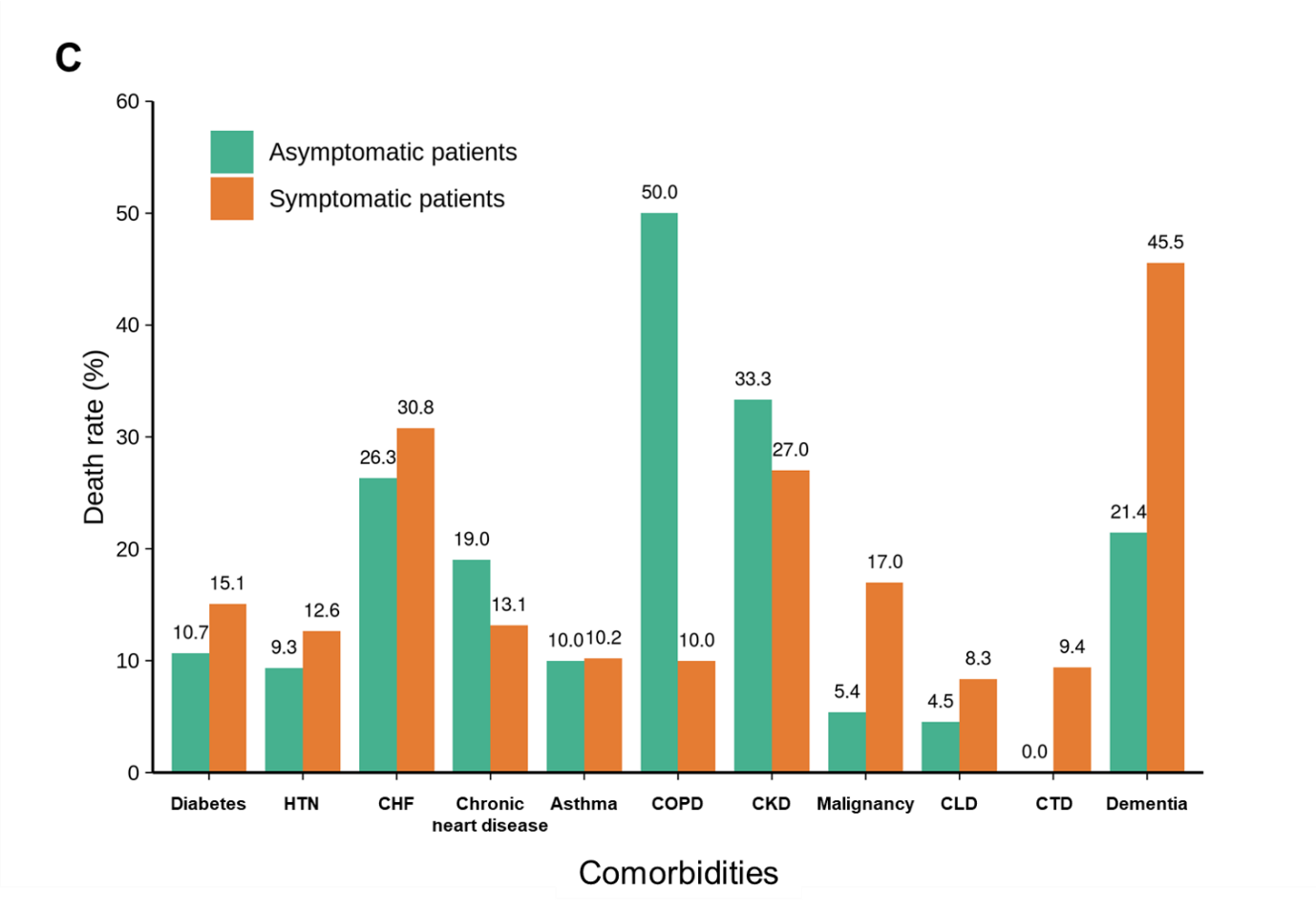

Appendix Figure 3. Calibration curves of the nomogram predicting 14-day (A), and 28-day (B) overall survival in asymptomatic patients with COVID-19. COVID-19, coronavirus disease 2019.

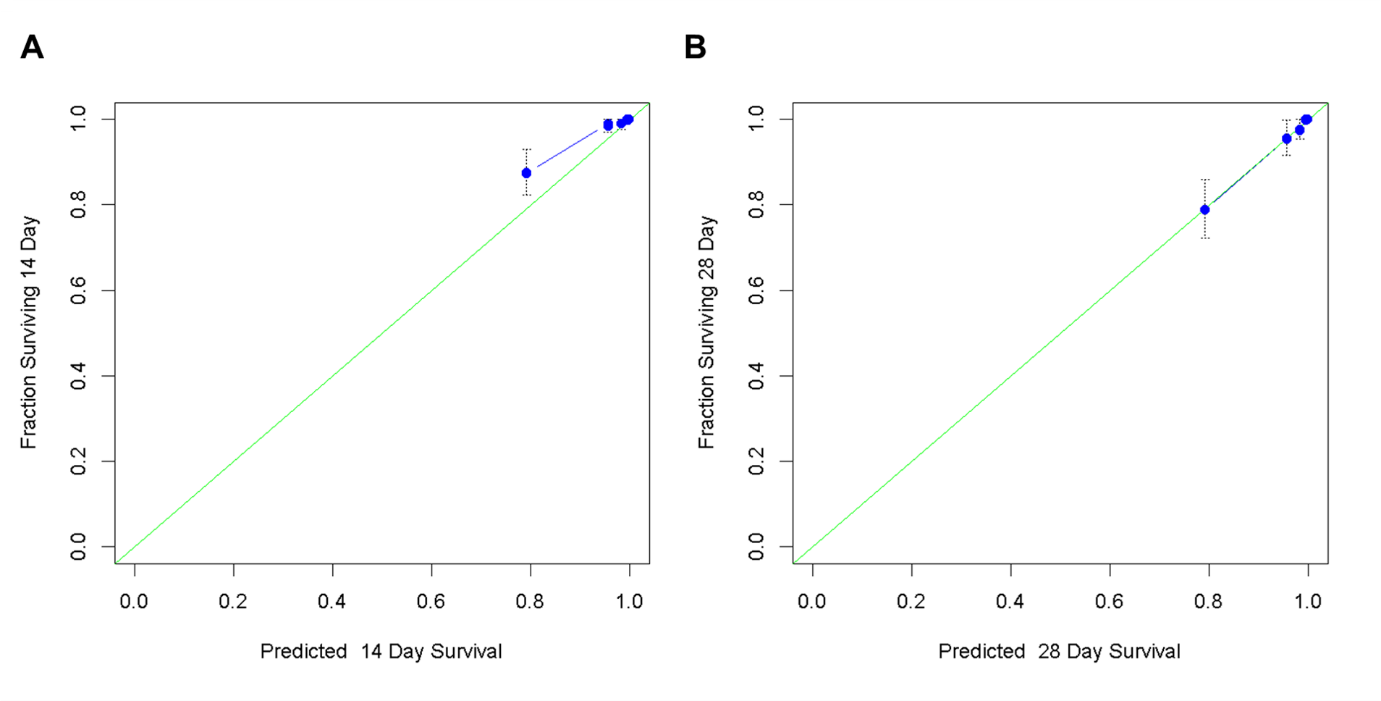

Appendix Table 1. Univariate and multivariate logistic regression analysis of predictors for admission to ICU in the patients with COVID-19

|  | Univariate | | Multivariate | |
| --- | --- | --- | --- | --- |
|  | Odd ratio (95% CI) | P value | Odd ratio (95% CI) | *P* value |
| Age 50-69 years | 4.69 (2.88 - 7.65) | <0.001 |  |  |
| ≥70 years | 15.27 (9.45 - 24.68) | <0.001 |  |  |
| Male | 2.34 (1.74 - 3.16) | <0.001 | 2.45 (1.63 - 3.67) | <0.001 |
| BMI <18.5 kg/m^2^ | 0.92 (0.45 - 1.90) | 0.82 | 0.85 (0.37 - 1.98) | 0.71 |
| Systolic BP <120 mmHg | 0.76 (0.53 - 1.10) | 0.15 |  |  |
| Diastolic BP <80 mmHg | 1.31 (0.98 - 1.76) | 0.07 | 1.25 (0.84 - 1.86) | 0.26 |
| Heart rate ≥100 /min | 1.89 (1.36 - 2.63) | <0.001 | 1.40 (0.88 - 2.22) | 0.16 |
| Body temperature ≥37.5℃ | 3.63 (2.68 - 4.92) | <0.001 | 1.3 (0.68 - 2.74) | 0.38 |
| CCIS ≥3 | 8.28 (5.68 - 12.05) | <0.001 | 3.63 (2.24 - 5.88) | <0.001 |
| Any symptom | 4.12 (2.42 - 7.00) | <0.001 |  |  |
| Febrile sense | 3.52 (2.62 - 4.72) | <0.001 | 1.92 (0.99 - 3.74) | 0.06 |
| Fatigue | 2.41 (1.44 - 4.05) | <0.001 | 1.17 (0.59 - 2.32) | 0.66 |
| Dyspnea | 9.00 (6.67 - 12.14) | <0.001 | 4.65 (3.13 - 6.90) | <0.001 |
| Altered mentality | 16.01 (7.60 - 33.73) | <0.001 | 6.06 (1.56 - 23.50) | 0.009 |
| Hemoglobin <12.0 g/dl | 2.36 (1.72 - 3.26) | <0.001 | 2.14 (1.38 - 3.31) | <0.001 |
| Lymphocyte counts <800 /μl | 6.68 (4.83 - 9.25) | <0.001 | 1.70 (1.08 - 2.65) | 0.02 |
| Platelet counts <150,000 /μl | 3.05 (2.16 - 4.31) | <0.001 | 1.34 (0.84 - 2.14) | 0.22 |

BMI, body mass index; BP, blood pressure; CCIS, age-adjusted Charlson comorbidity index score; CI, confidence interval; COPD, chronic obstructive pulmonary disease; COVID-19, coronavirus disease 2019; ICU, intensive care unit; OR, odd ratio.
